## Supplemental Table S2 for "Inferring personal intake recommendations of phosphorous and potassium for end-stage renal failure patients by simulating with Bayesian hierarchical multivariate model"

**Table S2: Strongest correlations between all personal effects and effects of potassium and phosphorous. This estimated structure of correlations is used to infer the personal effects based on the intake and the corresponding concentrations.**

| Personal effects of nutrients | | |
|---|---|---|
| **Effect 1** | **Effect 2** | **Correlation** |
| Sodium -> fP-Pi | Potassium-> P-Alb | 0.049 |
| Gender -> P-Alb | Potassium -> P-K | 0.046 |
| Fiber -> P-K | Potassium -> fP-Pi | 0.044 |
| PUFA, E% -> P-K | Potassium -> P-K | 0.043 |
| P-Alb level | Potassium-> P-Alb | 0.041 |
| Sodium -> P-Alb | Potassium -> fP-Pi | 0.037 |
| Blood lipid medication -> P-K | Potassium -> P-K | 0.035 |
| Salt -> P-Alb | Potassium -> P-K | 0.034 |
| P-Alb level | Potassium -> fP-Pi | 0.032 |
| SFA, E% -> P-K | Potassium-> P-Alb | 0.031 |
| Water -> P-K | Potassium -> fP-Pi | -0.030 |
| Water -> fP-Pi | Potassium -> fP-Pi | -0.030 |
| Hydroxycholecalciferol -> fP-Pi | Potassium -> fP-Pi | -0.031 |
| MUFA, E% -> P-K | Potassium -> P-K | -0.033 |
| Phosphorous -> P-K | Potassium -> P-K | -0.035 |
| Salt -> P-K | Potassium-> P-Alb | -0.040 |
| Diabetes medication -> P-Alb | Potassium-> P-Alb | -0.040 |
| Water -> P-Alb | Potassium -> P-K | -0.046 |
| Calcium -> fP-Pi | Potassium -> P-K | -0.051 |
| Phosphorous -> fP-Pi | Potassium -> P-K | -0.058 |
| fP-Pi level | Phosphorous -> P-K | 0.088 |
| Renavit -> P-Alb | Phosphorous -> P-K | 0.070 |
| Diabetes medication -> P-K | Phosphorous -> fP-Pi | 0.056 |
| Hydroxycholecalciferol -> fP-Pi | Phosphorous -> P-K | 0.053 |
| SFA, E% -> fP-Pi | Phosphorous-> P-Alb | 0.051 |
| Vitamin D -> P-Alb | Phosphorous -> P-K | 0.051 |
| Protein, g/kg -> fP-Pi | Phosphorous -> P-K | 0.049 |
| Hydroxycholecalciferol -> P-K | Phosphorous -> fP-Pi | 0.041 |
| Diabetes medication -> fP-Pi | Phosphorous -> P-K | 0.039 |
| MUFA, E% -> P-Alb | Phosphorous -> P-K | 0.036 |
| P-K level | Phosphorous-> P-Alb | -0.037 |
| Blood lipid medication -> P-Alb | Phosphorous-> P-Alb | -0.037 |
| MUFA, E% -> P-K | Phosphorous-> P-Alb | -0.039 |
| Calcium -> P-K | Phosphorous -> fP-Pi | -0.044 |
| Hydroxycholecalciferol -> P-K | Phosphorous -> P-K | -0.050 |
| Fiber -> P-Alb | Phosphorous-> P-Alb | -0.051 |
| Fat E% -> fP-Pi | Phosphorous -> fP-Pi | -0.053 |
| Water -> P-K | Phosphorous -> P-K | -0.055 |
| Diabetes medication -> P-Alb | Phosphorous -> P-K | -0.058 |
| Carbonhydrates, E% -> P-K | Phosphorous -> fP-Pi | -0.069 |
