## Supplemental Table S3 for "Inferring personal intake recommendations of phosphorous and potassium for end-stage renal failure patients by simulating with Bayesian hierarchical multivariate model"

| Nutrient | Conc. | General effect | Home hemodialysis | | | Hospital hemodialysis | | | Peritoneal dialysis | | |
|---|---|---|---|---|---|---|---|---|---|---|---|
| | | | **avg** | **min** | **max** | **avg** | **min** | **max** | **avg** | **min** | **max** |
| Water | P-Alb | -2.04 [−6.16; 2.97] | **-2.07** [−5.76; 1.76] | -2.60 [−8.07; 2.01] | -2.06 [−6.13; 2.47] | **-0.23** [−3.21; 2.52] | -0.60 [−4.28; 2.84] | 0.11 [−3.60; 3.77] | **-1.19** [−5.93; 4.22] | -1.53 [−6.39; 4.00] | -1.12 [−5.86; 4.42] |
| Blood lipid medication | P-Alb | 0.36 [−6.33; 7.76] | **1.25** [−6.00; 10.22] | 0.24 [−8.77; 11.01] | 1.97 [−6.60; 12.07] | **0.14** [−5.39; 5.61] | -1.17 [−7.39; 5.64] | 1.19 [−4.35; 6.96] | **1.14** [−5.92; 10.13] | 0.60 [−8.54; 9.56] | 1.81 [−5.82; 11.07] |
| Hydroxycholecalciferol | P-K | 0.50 [−3.57; 3.68] | **-0.42** [−5.08; 2.69] | -0.52 [−5.31; 2.40] | -0.19 [−5.07; 2.73] | **-0.51** [−2.17; 1.19] | -0.82 [−2.69; 1.19] | -0.25 [−2.28; 2.02] | **-0.94** [−3.42; 1.72] | -1.08 [−3.85; 1.76] | -0.83 [−3.48; 1.84] |
| Calcium | P-Alb | -0.08 [−10.35; 6.03] | **1.85** [−2.93; 7.46] | 1.22 [−3.36; 6.27] | 2.25 [−3.61; 9.68] | **0.88** [−3.47; 4.95] | -0.32 [−5.36; 4.17] | 1.59 [−3.38; 6.13] | **1.64** [−2.90; 6.84] | 2.03 [−3.76; 8.83] | 2.60 [−3.21; 11.67] |
| Sodium | P-Alb | 1.22 [−8.68; 15.01] | **2.23** [−5.69; 12.83] | 2.00 [−6.13; 11.84] | 2.80 [−5.80; 15.93] | **-0.87** [−6.51; 5.45] | -1.39 [−7.65; 5.06] | -0.28 [−6.33; 7.10] | **-3.02** [−13.00; 7.48] | -3.28 [−13.36; 7.35] | -2.60 [−12.82; 8.02] |
| Salt | P-Alb | 1.60 [−8.59; 10.81] | **3.76** [−7.68; 12.53] | 3.28 [−8.23; 11.96] | 4.52 [−6.23; 12.73] | **1.82** [−3.68; 6.93] | 1.27 [−4.49; 6.56] | 2.27 [−4.09; 9.07] | **-0.39** [−11.05; 8.80] | -0.81 [−12.25; 8.04] | -0.31 [−11.56; 8.59] |
| Gender | P-Alb | 1.67 [−5.48; 10.14] | **0.36** [−6.76; 6.92] | -0.53 [−11.15; 7.42] | 1.69 [−8.20; 10.28] | **2.73** [−3.79; 9.68] | 1.43 [−7.72; 9.22] | 4.13 [−3.80; 11.67] | **1.64** [−7.19; 9.96] | 0.77 [−11.13; 10.90] | 2.73 [−7.01; 13.42] |
| Phosphate binder med. | P-Alb | -2.78 [−9.87; 4.87] | **-2.68** [−10.50; 7.83] | -4.26 [−18.77; 8.61] | -1.59 [−9.86; 8.98] | **-1.95** [−9.93; 6.69] | -4.25 [−16.99; 7.20] | -0.96 [−11.15; 10.81] | **-0.37** [−8.93; 8.19] | -1.32 [−10.67; 7.77] | 0.84 [−10.26; 12.01] |
| Energy, kcal/kg | P-Alb | 1.60 [−11.49; 15.39] | **-0.40** [−12.86; 13.00] | -1.39 [−13.20; 11.42] | 0.69 [−12.25; 15.89] | **-1.40** [−12.93; 9.31] | -2.93 [−14.33; 8.17] | -0.18 [−12.79; 11.23] | **1.06** [−11.93; 16.56] | -0.18 [−13.30; 15.81] | 2.15 [−12.02; 18.26] |
| Carbohydrates, E% | P-Alb | 1.29 [−14.77; 15.35] | **2.67** [−13.38; 18.00] | 2.28 [−13.04; 17.08] | 3.71 [−11.99; 18.43] | **1.80** [−13.38; 15.62] | 0.91 [−15.21; 15.17] | 2.33 [−11.97; 15.69] | **0.82** [−15.89; 15.37] | 0.16 [−16.99; 15.88] | 1.33 [−15.87; 15.52] |
| Monounsaturated Fatty Acids, E% | P-Alb | 1.81 [−6.77; 13.45] | **2.82** [−3.72; 12.69] | 1.80 [−5.70; 12.61] | 3.92 [−3.14; 13.42] | **1.39** [−4.01; 6.82] | 0.71 [−5.18; 6.70] | 2.13 [−3.10; 7.39] | **0.38** [−7.41; 8.35] | -0.65 [−9.54; 8.54] | 0.87 [−6.41; 8.64] |
| Saturated Fatty Acids, E% | P-Alb | -2.58 [−9.85; 4.58] | **-3.39** [−10.30; 3.85] | -4.07 [−11.26; 3.42] | -2.80 [−9.88; 5.06] | **-2.23** [−9.00; 4.21] | -2.80 [−9.46; 4.27] | -1.17 [−7.44; 4.97] | **-2.19** [−9.54; 5.17] | -2.48 [−9.24; 5.09] | -1.63 [−9.61; 5.69] |
| Hydroxycholecalciferol | P-Alb | 1.71 [−6.25; 10.13] | **2.12** [−5.96; 12.13] | 1.61 [−6.54; 10.42] | 3.09 [−5.98; 13.35] | **0.90** [−5.30; 7.19] | 0.54 [−6.56; 8.47] | 1.68 [−6.08; 9.36] | **3.16** [−6.04; 13.07] | 2.24 [−7.66; 12.55] | 3.47 [−6.70; 13.94] |
| Polyunsaturated Fatty Acids, E% | P-Alb | -0.93 [−6.18; 5.86] | **-1.21** [−5.89; 3.58] | -1.43 [−6.40; 4.03] | -0.89 [−5.79; 4.35] | **-2.05** [−5.86; 1.40] | -2.42 [−6.01; 1.27] | -1.64 [−5.96; 2.41] | **-1.17** [−5.52; 3.44] | -1.27 [−6.31; 3.49] | -0.92 [−5.68; 3.78] |
| Potassium | P-Alb | -3.14 [−15.40; 4.12] | **-3.89** [−9.99; 1.30] | -5.04 [−12.20; 0.79] | -3.63 [−9.67; 2.06] | **-1.06** [−4.53; 1.96] | -1.97 [−8.76; 2.01] | 0.34 [−3.64; 3.96] | **-0.24** [−5.53; 5.54] | -1.02 [−7.03; 5.17] | 0.27 [−5.92; 8.05] |
| Renavit | P-Alb | 10.04 [−12.10; 24.48] | **11.73** [−8.72; 25.68] | 10.55 [−8.97; 24.74] | 12.77 [−7.93; 28.05] | **9.33** [−9.77; 24.03] | 7.75 [−10.19; 22.86] | 10.62 [−10.79; 25.61] | **8.25** [−12.70; 22.49] | 7.28 [−12.78; 21.69] | 9.12 [−12.99; 23.78] |
| Fat E% | P-Alb | 3.19 [−14.81; 19.59] | **2.32** [−16.46; 20.22] | 1.52 [−17.42; 20.12] | 3.00 [−15.81; 20.48] | **3.40** [−12.94; 18.61] | 2.52 [−15.43; 17.92] | 4.14 [−12.29; 19.78] | **3.50** [−14.06; 19.88] | 2.88 [−14.63; 19.85] | 3.95 [−14.63; 21.22] |
| Phosphorous | P-Alb | -0.19 [−7.16; 6.56] | **-2.10** [−10.73; 5.28] | -3.09 [−12.68; 4.94] | -1.87 [−12.44; 7.81] | **0.27** [−5.57; 5.95] | -0.41 [−6.77; 6.08] | 1.73 [−4.98; 8.63] | **0.80** [−7.31; 8.48] | 0.10 [−8.68; 8.11] | 1.22 [−6.42; 8.81] |
| Protein, g/kg | P-Alb | 0.33 [−12.64; 13.11] | **-1.35** [−16.07; 11.86] | -2.72 [−17.16; 10.61] | -0.19 [−15.66; 14.04] | **-0.28** [−11.68; 11.80] | -0.95 [−12.82; 11.39] | 0.98 [−11.00; 15.10] | **1.09** [−13.08; 14.12] | 0.74 [−12.95; 13.72] | 1.62 [−12.93; 15.03] |
| Vitamin D | P-Alb | 1.60 [−1.28; 4.01] | **1.55** [−1.01; 4.27] | 0.79 [−3.52; 4.84] | 2.39 [−0.82; 6.41] | **0.79** [−1.25; 2.87] | 0.11 [−3.45; 3.32] | 1.45 [−1.76; 4.78] | **1.50** [−1.49; 4.58] | 1.24 [−2.88; 5.29] | 2.22 [−1.24; 6.60] |
| Fiber | P-Alb | 1.19 [−5.02; 7.55] | **1.43** [−2.36; 5.60] | 1.00 [−3.40; 6.23] | 2.02 [−2.56; 7.51] | **1.34** [−1.91; 4.70] | 0.70 [−3.35; 4.61] | 2.01 [−2.06; 6.49] | **1.60** [−3.85; 6.61] | 1.28 [−4.24; 6.85] | 1.92 [−4.02; 7.59] |
| Diabetes medication | P-Alb | 0.67 [−6.75; 8.48] | **0.45** [−6.07; 8.60] | -0.22 [−8.31; 8.33] | 1.58 [−5.59; 9.25] | **-0.12** [−6.22; 5.63] | -0.98 [−7.54; 5.88] | 1.55 [−6.13; 11.84] | **0.49** [−7.22; 8.87] | 0.03 [−9.63; 9.68] | 1.40 [−8.00; 11.06] |
| Phosphate binder med. | P-K | 0.28 [−1.55; 2.75] | **0.25** [−1.83; 2.88] | -0.21 [−2.39; 2.53] | 0.61 [−2.15; 3.88] | **0.40** [−1.23; 2.46] | 0.04 [−2.01; 2.50] | 0.68 [−1.49; 3.63] | **0.26** [−1.45; 2.40] | -0.14 [−2.83; 2.70] | 0.70 [−1.44; 3.12] |
| Phosphate binder med. | fP-Pi | 0.50 [−1.50; 2.94] | **0.61** [−0.97; 2.71] | 0.33 [−2.11; 2.69] | 0.82 [−1.44; 3.28] | **0.41** [−0.90; 2.00] | 0.15 [−1.86; 1.99] | 0.73 [−1.34; 3.02] | **0.12** [−1.71; 1.81] | -0.05 [−2.23; 1.87] | 0.18 [−1.91; 2.04] |
| Protein, E% | P-Alb | 0.64 [−7.30; 7.41] | **2.08** [−4.84; 8.78] | 1.70 [−5.21; 8.26] | 2.67 [−4.80; 10.24] | **1.72** [−5.00; 8.28] | 1.17 [−5.17; 7.86] | 2.40 [−5.78; 9.59] | **1.27** [−5.70; 8.44] | 0.88 [−7.16; 8.59] | 1.78 [−5.68; 8.87] |
| Protein, g/kg | P-K | -0.22 [−2.93; 2.74] | **0.09** [−2.42; 2.57] | -0.06 [−2.63; 2.54] | 0.21 [−3.09; 2.96] | **0.03** [−2.50; 2.41] | -0.22 [−3.16; 2.65] | 0.18 [−2.37; 2.65] | **-0.33** [−3.63; 2.99] | -0.36 [−3.95; 3.03] | -0.25 [−3.63; 3.12] |
| Carbohydrates, E% | P-K | 1.05 [−2.32; 4.69] | **1.26** [−2.36; 4.61] | 1.12 [−2.64; 4.81] | 1.32 [−2.46; 4.82] | **0.85** [−2.16; 3.43] | 0.67 [−2.37; 3.40] | 0.93 [−2.40; 3.47] | **0.74** [−2.67; 4.07] | 0.68 [−2.99; 4.04] | 0.83 [−2.73; 4.47] |
| Calcium | P-K | -0.09 [−1.49; 1.54] | **-0.46** [−2.59; 0.97] | -0.63 [−3.83; 1.03] | -0.36 [−2.93; 1.41] | **0.01** [−0.94; 1.17] | -0.15 [−1.92; 1.29] | 0.38 [−0.76; 2.19] | **-0.13** [−1.62; 1.29] | -0.37 [−2.41; 1.41] | 0.05 [−1.49; 1.56] |
| Energy, kcal/kg | P-K | 0.30 [−2.53; 4.98] | **-0.19** [−2.92; 2.96] | -0.30 [−3.37; 2.87] | -0.11 [−2.54; 2.94] | **-0.14** [−2.52; 2.41] | -0.24 [−2.71; 2.37] | -0.09 [−2.64; 2.53] | **0.29** [−3.70; 4.70] | 0.17 [−3.81; 4.57] | 0.43 [−3.70; 4.95] |
| Salt | P-K | 0.11 [−1.82; 2.50] | **0.93** [−1.37; 2.94] | 0.74 [−1.96; 2.72] | 1.09 [−1.35; 3.42] | **0.18** [−0.79; 1.10] | -0.09 [−2.57; 1.23] | 0.38 [−0.64; 1.41] | **-0.24** [−2.89; 1.63] | -0.50 [−3.94; 1.64] | -0.20 [−3.15; 1.68] |
| Gender | P-K | -0.04 [−2.53; 2.63] | **0.41** [−1.36; 2.47] | 0.35 [−1.54; 2.50] | 0.57 [−1.36; 2.75] | **0.26** [−1.17; 1.78] | 0.07 [−1.57; 1.95] | 0.39 [−1.40; 2.49] | **0.92** [−1.97; 4.50] | 0.76 [−2.39; 4.48] | 1.07 [−1.90; 4.28] |
| Blood lipid medication | P-K | -0.65 [−3.47; 1.82] | **0.11** [−2.42; 4.45] | -0.21 [−3.88; 4.27] | 0.36 [−2.46; 5.41] | **-0.32** [−1.68; 0.98] | -0.61 [−2.45; 0.86] | 0.10 [−1.30; 1.86] | **-0.55** [−3.45; 1.73] | -0.96 [−5.15; 1.91] | -0.23 [−3.25; 3.33] |
