## Supplemental Table S3 continues for "Inferring personal intake recommendations of phosphorous and potassium for end-stage renal failure patients by simulating with Bayesian hierarchical multivariate model"

| Nutrient | Conc. | General effect | Home hemodialysis | | | Hospital hemodialysis | | | Peritoneal dialysis | | |
|---|---|---|---|---|---|---|---|---|---|---|---|
| | | | avg | min | max | avg | min | max | avg | min | max |
| Carbohydrates, E% | fP-Pi | 0.29 [−2.56; 2.87] | **0.00** [−1.88; 1.73] | -0.07 [−1.88; 1.68] | 0.03 [−1.82; 1.79] | **0.16** [−1.40; 1.58] | 0.08 [−1.66; 1.53] | 0.27 [−1.08; 1.59] | **0.06** [−1.90; 1.87] | 0.01 [−1.89; 1.85] | 0.08 [−1.99; 1.97] |
| Monounsaturated Fatty Acids, E% | P-K | 0.43 [−1.45; 3.02] | **0.76** [−0.85; 3.37] | 0.72 [−0.93; 3.46] | 0.93 [−0.87; 3.74] | **0.52** [−0.75; 2.02] | 0.28 [−1.18; 1.77] | 0.66 [−0.96; 3.02] | **0.58** [−1.48; 2.81] | 0.45 [−1.80; 2.80] | 0.72 [−1.20; 2.68] |
| Sodium | fP-Pi | 0.19 [−1.69; 1.93] | **0.63** [−0.91; 3.46] | 0.57 [−0.85; 3.26] | 0.70 [−0.86; 3.47] | **0.12** [−0.65; 0.72] | 0.05 [−0.76; 0.68] | 0.20 [−0.57; 0.85] | **0.41** [−1.65; 3.99] | 0.36 [−1.60; 3.80] | 0.47 [−1.68; 4.15] |
| Sodium | P-K | -0.15 [−1.98; 2.35] | **-0.06** [−1.88; 2.31] | -0.15 [−1.98; 2.23] | -0.02 [−1.88; 2.32] | **-0.25** [−1.74; 1.63] | -0.40 [−1.86; 1.45] | -0.05 [−1.76; 2.26] | **-0.54** [−2.64; 2.16] | -0.60 [−2.74; 2.22] | -0.39 [−2.55; 2.47] |
| Potassium | P-K | -0.46 [−3.35; 2.02] | **-0.06** [−1.48; 1.55] | -0.32 [−2.14; 1.32] | 0.14 [−1.57; 1.76] | **0.14** [−0.58; 1.02] | -0.11 [−1.40; 1.27] | 0.50 [−0.52; 1.64] | **-0.21** [−1.66; 0.98] | -0.51 [−2.22; 1.17] | 0.16 [−1.50; 2.45] |
| Diabetes medication | fP-Pi | 0.20 [−0.64; 1.86] | **0.19** [−0.71; 1.33] | 0.04 [−1.14; 1.37] | 0.37 [−0.74; 1.72] | **0.05** [−0.68; 0.77] | -0.09 [−1.12; 0.90] | 0.10 [−0.76; 1.09] | **0.17** [−0.95; 1.93] | 0.04 [−1.28; 2.04] | 0.33 [−1.12; 2.12] |
| Polyunsaturated Fatty Acids, E% | fP-Pi | 0.22 [−1.14; 2.26] | **-0.52** [−1.63; 0.31] | -0.53 [−1.81; 0.37] | -0.50 [−1.62; 0.40] | **-0.34** [−0.89; 0.09] | -0.40 [−1.04; 0.10] | -0.29 [−0.80; 0.16] | **-0.25** [−0.97; 0.62] | -0.28 [−1.06; 0.64] | -0.21 [−0.93; 0.64] |
| Blood lipid medication | fP-Pi | 0.02 [−1.88; 3.27] | **-0.81** [−3.36; 1.66] | -0.95 [−3.36; 2.11] | -0.67 [−3.58; 1.79] | **-0.51** [−1.24; 0.19] | -0.82 [−2.45; 0.29] | -0.33 [−1.19; 0.49] | **-0.14** [−1.96; 2.11] | -0.33 [−2.22; 1.70] | -0.07 [−2.23; 2.54] |
| Diabetes medication | P-K | 5.41 [−1.85; 23.43] | **-0.17** [−1.92; 2.00] | -0.35 [−2.73; 2.46] | 0.07 [−1.90; 2.35] | **-0.13** [−1.29; 1.14] | -0.31 [−2.35; 1.47] | 0.08 [−1.18; 1.81] | **-0.65** [−4.90; 2.38] | -0.76 [−4.82; 2.23] | -0.54 [−5.11; 2.94] |
| Energy, kcal/kg | fP-Pi | 0.50 [−1.50; 2.17] | **0.58** [−0.90; 2.05] | 0.46 [−0.95; 1.93] | 0.68 [−0.90; 2.25] | **0.71** [−0.72; 1.96] | 0.63 [−0.76; 1.91] | 0.80 [−0.72; 2.08] | **0.51** [−1.79; 2.22] | 0.45 [−1.83; 2.21] | 0.56 [−1.88; 2.41] |
| Fat E% | fP-Pi | 0.21 [−1.81; 2.17] | **0.51** [−1.48; 2.46] | 0.46 [−1.50; 2.43] | 0.57 [−1.46; 2.74] | **0.57** [−1.22; 2.58] | 0.50 [−1.26; 2.55] | 0.63 [−1.25; 2.79] | **0.18** [−1.91; 2.15] | 0.13 [−1.82; 2.12] | 0.24 [−2.00; 2.17] |
| Renavit | P-K | 0.94 [−3.25; 7.50] | **0.29** [−3.83; 6.53] | 0.11 [−4.16; 6.46] | 0.58 [−3.46; 6.62] | **0.76** [−2.53; 5.74] | 0.49 [−3.01; 5.34] | 0.93 [−2.38; 5.89] | **0.88** [−3.09; 6.47] | 0.92 [−3.27; 6.59] | 1.08 [−2.84; 6.36] |
| Water | P-K | 0.16 [−0.99; 1.79] | **0.18** [−0.59; 1.08] | -0.04 [−1.81; 1.35] | 0.36 [−0.54; 1.31] | **-0.24** [−0.92; 0.44] | -0.48 [−1.67; 0.65] | 0.01 [−0.69; 0.56] | **-0.10** [−1.53; 1.13] | -0.33 [−1.93; 1.27] | 0.01 [−1.53; 1.54] |
| Water | fP-Pi | 0.11 [−0.71; 0.73] | **0.14** [−0.29; 0.60] | 0.03 [−0.88; 0.69] | 0.24 [−0.37; 0.89] | **0.17** [−0.13; 0.55] | 0.00 [−0.30; 0.34] | 0.23 [−0.20; 0.87] | **0.05** [−0.81; 0.99] | -0.08 [−1.18; 0.97] | 0.21 [−0.69; 1.08] |
| Fiber | P-K | -0.92 [−6.53; 1.42] | **0.23** [−0.72; 1.48] | -0.07 [−1.45; 1.17] | 0.46 [−0.85; 1.85] | **0.03** [−0.93; 0.94] | -0.29 [−1.45; 0.96] | 0.28 [−1.04; 1.33] | **0.47** [−1.04; 2.48] | 0.31 [−1.32; 2.59] | 0.62 [−1.21; 2.79] |
| Renavit | fP-Pi | 0.14 [−2.13; 3.06] | **0.36** [−2.03; 2.93] | 0.28 [−2.04; 2.94] | 0.39 [−2.03; 2.96] | **0.20** [−1.60; 1.78] | 0.14 [−1.73; 1.80] | 0.26 [−1.63; 1.89] | **0.77** [−2.49; 4.08] | 0.70 [−2.59; 3.99] | 0.83 [−2.47; 4.02] |
| Vitamin D | P-K | -0.17 [−1.20; 1.11] | **-0.15** [−0.94; 0.60] | -0.25 [−1.23; 0.78] | -0.11 [−0.91; 0.79] | **-0.18** [−0.65; 0.45] | -0.33 [−1.04; 0.46] | 0.02 [−0.44; 0.60] | **-0.31** [−1.24; 0.72] | -0.49 [−1.32; 0.42] | -0.22 [−1.21; 0.95] |
| Protein, g/kg | fP-Pi | -0.80 [−2.41; 1.10] | **-0.84** [−2.37; 0.91] | -0.91 [−2.49; 0.86] | -0.78 [−2.52; 0.99] | **-0.81** [−2.03; 0.65] | -0.86 [−2.16; 0.57] | -0.74 [−2.03; 0.74] | **-0.64** [−2.32; 1.06] | -0.65 [−2.29; 1.10] | -0.60 [−2.38; 1.12] |
| Fat E% | P-K | -0.38 [−4.54; 3.29] | **-0.14** [−4.80; 3.86] | -0.26 [−5.16; 3.77] | -0.06 [−4.85; 4.02] | **-0.25** [−4.41; 3.08] | -0.40 [−4.67; 2.98] | -0.01 [−4.12; 3.17] | **-0.51** [−5.07; 3.44] | -0.65 [−5.50; 3.48] | -0.41 [−4.82; 3.39] |
| Protein, E% | fP-Pi | 0.20 [−1.06; 1.87] | **0.12** [−0.65; 0.79] | 0.08 [−0.70; 0.85] | 0.16 [−0.68; 0.81] | **0.24** [−0.49; 0.84] | 0.20 [−0.56; 0.87] | 0.30 [−0.39; 0.92] | **0.14** [−1.03; 1.09] | 0.09 [−1.04; 1.05] | 0.17 [−1.05; 1.16] |
| Monounsaturated Fatty Acids, E% | fP-Pi | -0.10 [−1.84; 1.77] | **0.06** [−0.73; 1.05] | 0.03 [−0.80; 1.06] | 0.12 [−0.77; 1.21] | **-0.13** [−0.89; 0.62] | -0.16 [−1.04; 0.63] | -0.05 [−0.88; 0.72] | **-0.02** [−0.95; 0.94] | -0.06 [−1.01; 0.97] | 0.02 [−0.91; 1.00] |
| Polyunsaturated Fatty Acids, E% | P-K | 0.29 [−1.25; 1.84] | **0.28** [−1.11; 1.73] | 0.21 [−1.28; 1.85] | 0.38 [−1.21; 2.24] | **0.15** [−0.71; 1.10] | 0.00 [−1.02; 1.07] | 0.25 [−0.76; 1.48] | **0.32** [−0.91; 1.86] | 0.19 [−1.05; 1.44] | 0.45 [−0.81; 2.22] |
| Phosphorous | P-K | 0.39 [−2.48; 2.31] | **0.25** [−1.41; 2.02] | 0.15 [−1.57; 1.79] | 0.36 [−1.54; 2.84] | **0.21** [−1.20; 1.69] | 0.10 [−1.37; 1.54] | 0.37 [−1.30; 2.61] | **0.51** [−1.42; 2.55] | 0.45 [−1.44; 2.39] | 0.59 [−1.49; 2.61] |
| Protein, E% | P-K | 0.49 [−1.46; 3.27] | **0.23** [−0.93; 2.24] | 0.13 [−1.23; 2.20] | 0.25 [−0.99; 2.24] | **0.28** [−1.23; 2.16] | 0.15 [−1.46; 2.25] | 0.42 [−1.03; 2.26] | **0.54** [−1.25; 3.15] | 0.46 [−1.49; 3.14] | 0.52 [−1.32; 3.09] |
| Saturated Fatty Acids, E% | P-K | 1.01 [−1.10; 3.03] | **0.86** [−1.48; 3.10] | 0.77 [−1.34; 2.93] | 0.95 [−1.50; 3.18] | **0.82** [−0.95; 2.82] | 0.64 [−1.47; 2.69] | 0.95 [−0.86; 3.08] | **1.06** [−1.01; 3.19] | 1.01 [−1.17; 3.28] | 1.18 [−0.85; 3.25] |
| Saturated Fatty Acids, E% | fP-Pi | -0.36 [−1.51; 0.79] | **-0.53** [−2.02; 0.56] | -0.58 [−2.04; 0.57] | -0.49 [−2.07; 0.67] | **-0.32** [−1.22; 0.50] | -0.39 [−1.34; 0.50] | -0.27 [−1.24; 0.65] | **-0.05** [−1.14; 0.96] | -0.06 [−1.19; 0.98] | -0.01 [−1.15; 1.03] |
| Fiber | fP-Pi | -0.21 [−1.69; 1.06] | **-0.11** [−0.60; 0.37] | -0.18 [−0.79; 0.36] | -0.08 [−0.56; 0.41] | **-0.11** [−0.66; 0.30] | -0.19 [−0.79; 0.21] | -0.05 [−0.69; 0.56] | **-0.13** [−1.11; 0.96] | -0.16 [−1.25; 0.97] | -0.09 [−1.15; 1.09] |
| Calcium | fP-Pi | -0.25 [−1.24; 0.42] | **-0.04** [−0.54; 0.56] | -0.11 [−0.93; 0.69] | 0.17 [−0.63; 1.12] | **-0.15** [−0.59; 0.22] | -0.30 [−1.19; 0.29] | 0.01 [−0.56; 0.66] | **-0.06** [−0.96; 0.74] | -0.24 [−1.12; 0.84] | 0.00 [−1.04; 1.04] |
| Hydroxycholecalciferol | fP-Pi | 1.51 [−1.59; 7.33] | **0.52** [−2.27; 3.05] | 0.40 [−2.44; 3.08] | 0.70 [−2.10; 3.25] | **0.49** [−0.30; 1.38] | 0.40 [−0.64; 1.38] | 0.59 [−0.42; 1.77] | **0.17** [−2.04; 1.54] | -0.05 [−2.09; 1.56] | 0.26 [−2.08; 1.65] |
| Gender | fP-Pi | -0.13 [−1.66; 0.88] | **-0.02** [−1.32; 1.06] | -0.14 [−1.51; 1.14] | 0.06 [−1.46; 1.22] | **-0.14** [−1.12; 0.76] | -0.28 [−1.42; 0.95] | 0.05 [−1.04; 1.10] | **-0.04** [−1.34; 1.14] | -0.07 [−1.41; 1.22] | 0.02 [−1.53; 1.54] |
| Vitamin D | fP-Pi | 0.10 [−0.37; 0.54] | **0.06** [−0.41; 0.58] | -0.07 [−0.69; 0.46] | 0.13 [−0.54; 1.01] | **0.15** [−0.14; 0.41] | 0.05 [−0.44; 0.52] | 0.29 [−0.14; 0.71] | **0.14** [−0.40; 0.64] | 0.16 [−0.52; 0.77] | 0.22 [−0.35; 0.82] |
| Potassium | fP-Pi | 0.07 [−0.88; 0.88] | **0.00** [−0.72; 0.76] | -0.05 [−0.82; 0.77] | 0.04 [−0.72; 0.78] | **0.06** [−0.34; 0.52] | 0.03 [−0.51; 0.53] | 0.09 [−0.35; 0.56] | **0.17** [−0.56; 0.97] | 0.15 [−0.68; 1.03] | 0.21 [−0.48; 1.05] |
| Phosphorous | fP-Pi | 0.08 [−0.90; 1.41] | **0.15** [−1.00; 1.13] | 0.10 [−1.19; 1.23] | 0.21 [−1.14; 1.21] | **-0.02** [−0.79; 0.79] | -0.09 [−0.95; 0.81] | 0.08 [−0.63; 0.76] | **0.16** [−0.91; 1.43] | 0.05 [−1.22; 1.43] | 0.20 [−0.90; 1.45] |
| Salt | fP-Pi | 0.61 [−1.26; 3.65] | **-0.66** [−3.44; 0.90] | -0.71 [−3.64; 0.86] | -0.62 [−3.37; 0.96] | **-0.11** [−0.52; 0.39] | -0.16 [−0.91; 0.42] | -0.04 [−0.54; 0.56] | **-0.66** [−3.86; 1.21] | -0.73 [−4.08; 1.25] | -0.64 [−3.74; 1.22] |
