## Supplemental Table S4 for "Inferring personal intake recommendations of phosphorous and potassium for end-stage renal failure patients by simulating with Bayesian hierarchical multivariate model"

**Table S4: Notation summary of the inference method**

| Notation | Description |
|---|---|
| $b$ | Personal effect, in addition to treatment effect |
| $c$ | Constant for shifting the distribution |
| $g$ | Treatment effect, in addition to the general effect |
| $G_k$ | Personal graphical model for patient $k$ |
| $G_k^*$ | Personal graphical model with modified intake |
| $i$ | Observation index |
| $j$ | Predictor index (nutrient and medication) |
| $J$ | Number of predictors |
| $k$ | Patient index |
| $K$ | Number of patients |
| $l$ | Dialysis treatment type index |
| $L$ | Maximum number of treatments |
| $l_x$ | Quantile of intake random variable used in the estimation |
| $l_\beta$ | Quantile of nutrient effect used in the estimation |
| $m$ | Concentration index |
| $M$ | Number of considered concentrations |
| $n$ | Number of observations |
| $p$ | Number of predictors |
| $P_m^{max}$ | Maximum probability for reaching target for concentration $m$ |
| $P^{max}$ | Maximum probability for reaching all the concentration targets |
| $Q$ | Recommended nutrient |
| $Q_r^{min}$ | Lower limit of 95%-quantile value for recommended nutrient $r$ |
| $Q_r^{max}$ | Upper limit of 95%-quantile value for recommended nutrient $r$ |
| $r$ | Recommended nutrient index |
| $R$ | Number of recommended nutrients |
| $s$ | Recommendation algorithm sample index |
| $S$ | Number of samples to draw from the recommendation algorithm |
| $Y$ | Concentration level |
| $Y_m^l$ | Lower limit of concentration $m$ |
| $Y_m^u$ | Upper limit of concentration $m$ |
| $X$ | General predictor, intake level or personal information |
| $\mathbf{X}$ | $p$ x $n$ matrix of predictor observations |
| $Z$ | Personally varying predictor, intake level or personal information |
| $\mathbf{Z}$ | $p$ x $n$ matrix of predictor observations |
| $\alpha$ | Shape parameter of gamma distribution |
| $\beta_{jm}$ | General effect of predictor $j$ to concentration $m$ |
| $\mu_m$ | Expected value of concentration $m$ |
| $\rho_{mn}$ | Correlation between random variables $m$ and $n$ |
| $\otimes$ | Kronecker product |
