## Supplemental Table S1 for "Inferring personal intake recommendations of phosphorous and potassium for end-stage renal failure patients by simulating with Bayesian hierarchical multivariate model"

**Table S1: Strongest correlations between all treatment effects and treatment effects of potassium and phosphorous. This estimated structure of correlations is used to infer the treatment effects based on the intake and the corresponding concentrations.**

| Treatment-level effects of nutrients | | |
|---|---|---|
| **Effect 1** | **Effect 2** | **Correlation** |
| Renavit → fP-Pi | Potassium→ P-Alb | 0.074 |
| Gender → fP-Pi | Potassium→ P-Alb | 0.071 |
| Sodium → P-K | Potassium → fP-Pi | 0.065 |
| Protein, g/kg → P-Alb | Potassium→ P-Alb | 0.063 |
| Sodium → P-K | Potassium → P-K | 0.058 |
| Calcium → P-K | Potassium → P-K | 0.053 |
| MUFA, E% → P-K | Potassium → P-K | 0.052 |
| PUFA, E% → P-Alb | Potassium → P-K | 0.050 |
| Diabetes medication → P-Alb | Potassium → P-K | 0.047 |
| Vitamin D → fP-Pi | Potassium→ P-Alb | 0.045 |
| Blood lipid medication → fP-Pi | Potassium → fP-Pi | 0.044 |
| Phosphorous → P-Alb | Potassium→ P-Alb | 0.043 |
| PUFA, E% → fP-Pi | Potassium → P-K | -0.042 |
| Salt → P-K | Potassium→ P-Alb | -0.043 |
| Hydroxycholecalciferol → P-K | Potassium → P-K | -0.045 |
| Sodium → fP-Pi | Potassium→ P-Alb | -0.048 |
| Sodium → P-Alb | Potassium→ P-Alb | -0.049 |
| MUFA, E% → P-Alb | Potassium→ P-Alb | -0.051 |
| P-Alb level | Potassium → P-K | -0.056 |
| PUFA, E% → P-Alb | Potassium → fP-Pi | -0.059 |
| Potassium → P-K | Phosphorous → P-K | 0.066 |
| Blood lipid medication → P-K | Phosphorous → fP-Pi | 0.054 |
| Calcium → P-K | Phosphorous → fP-Pi | 0.051 |
| MUFA, E% → P-K | Phosphorous → P-K | 0.050 |
| Protein, E% → fP-Pi | Phosphorous → fP-Pi | 0.044 |
| Salt → fP-Pi | Phosphorous→ P-Alb | 0.043 |
| PUFA, E% → fP-Pi | Phosphorous→ P-Alb | 0.042 |
| Calcium → P-K | Phosphorous → P-K | 0.038 |
| SFA, E% → fP-Pi | Phosphorous → fP-Pi | 0.036 |
| Hydroxycholecalciferol → P-K | Phosphorous → fP-Pi | -0.037 |
| SFA, E% → fP-Pi | Phosphorous → P-K | -0.037 |
| Carbonhydrates, E% → P-K | Phosphorous → P-K | -0.039 |
| Phosphorous → fP-Pi | Phosphorous→ P-Alb | -0.039 |
| Salt → P-Alb | Phosphorous → fP-Pi | -0.039 |
| Carbonhydrates, E% → fP-Pi | Phosphorous → P-K | -0.040 |
| Phosphate binder med. → P-Alb | Phosphorous → fP-Pi | -0.042 |
| PUFA, E% → fP-Pi | Phosphorous → fP-Pi | -0.048 |
| SFA, E% → P-Alb | Phosphorous → fP-Pi | -0.053 |
| Carbonhydrates, E% → P-K | Phosphorous → fP-Pi | -0.060 |
| Calcium → fP-Pi | Phosphorous → P-K | -0.063 |
